# Automated Identification of Heart Failure with Reduced Ejection Fraction using Deep Learning-based Natural Language Processing

**DOI:** 10.1101/2023.09.10.23295315

**Authors:** Arash A. Nargesi, Philip Adejumo, Lovedeep Dhingra, Benjamin Rosand, Astrid Hengartner, Andreas Coppi, Simon Benigeri, Sounok Sen, Tariq Ahmad, Girish N Nadkarni, Zhenqiu Lin, Faraz S. Ahmad, Harlan M Krumholz, Rohan Khera

## Abstract

**Background:** The lack of automated tools for measuring care quality has limited the implementation of a national program to assess and improve guideline-directed care in heart failure with reduced ejection fraction (HFrEF). A key challenge for constructing such a tool has been an accurate, accessible approach for identifying patients with HFrEF at hospital discharge, an opportunity to evaluate and improve the quality of care.

**Methods:** We developed a novel deep learning-based language model for identifying patients with HFrEF from discharge summaries using a semi-supervised learning framework. For this purpose, hospitalizations with heart failure at Yale New Haven Hospital (YNHH) between 2015 to 2019 were labeled as HFrEF if the left ventricular ejection fraction was under 40% on antecedent echocardiography. The model was internally validated with model-based net reclassification improvement (NRI) assessed against chart-based diagnosis codes. We externally validated the model on discharge summaries from hospitalizations with heart failure at Northwestern Medicine, community hospitals of Yale New Haven Health in Connecticut and Rhode Island, and the publicly accessible MIMIC-III database, confirmed with chart abstraction.

**Results:** A total of 13,251 notes from 5,392 unique individuals (mean age 73 ± 14 years, 48% female), including 2,487 patients with HFrEF (46.1%), were used for model development (train/held-out test: 70/30%). The deep learning model achieved an area under receiving operating characteristic (AUROC) of 0.97 and an area under precision-recall curve (AUPRC) of 0.97 in detecting HFrEF on the held-out set. In external validation, the model had high performance in identifying HFrEF from discharge summaries with AUROC 0.94 and AUPRC 0.91 on 19,242 notes from Northwestern Medicine, AUROC 0.95 and AUPRC 0.96 on 139 manually abstracted notes from Yale community hospitals, and AUROC 0.91 and AUPRC 0.92 on 146 manually reviewed notes at MIMIC-III. Model-based prediction of HFrEF corresponded to an overall NRI of 60.2 ± 1.9% compared with the chart diagnosis codes (p-value < 0.001) and an increase in AUROC from 0.61 [95% CI: 060-0.63] to 0.91 [95% CI 0.90-0.92].

**Conclusions:** We developed and externally validated a deep learning language model that automatically identifies HFrEF from clinical notes with high precision and accuracy, representing a key element in automating quality assessment and improvement for individuals with HFrEF.

## BACKGROUND

Heart failure with reduced ejection fraction (HFrEF) accounts for nearly half of the hospitalizations for heart failure, with a nearly 50% mortality in 5 years after diagnosis (1). Several evidence-based therapies can improve survival and lower the risk of hospitalization in patients with HFrEF (2). However, only 1 in 5 eligible individuals receive all the first-line treatments for HFrEF in the United States (3,4). This gap highlights the urgent need for new strategies to implement guideline-directed therapies in clinical practice.

Clinical notes from electronic health records (EHR) are rich real-world data sources that can support systematic approaches to identify patient groups, including those with HFrEF, a linchpin for the assessment and improvement of care. Natural language processing (NLP) of clinical notes can automate the identification of individuals with HFrEF from the EHR (5,6), and has been recognized by the 2022 AHA/ACC/HFSA guideline for the management of heart failure as a novel implementation technique to assess the quality of care in patients with heart failure (7). NLP-based algorithms address the limitations of structured data, such as poorly recorded diagnosis codes and single-source echocardiography measurements. However, previous NLP-based models have been limited to semantic algorithms that search for pre-specified terms with unclear performance in identifying HFrEF (8,9). Misspelling of disease diagnosis, frequent use of abbreviations in clinical documentation, and variations in textual descriptions of clinical entities such as heart failure with recovered ejection fraction, are barriers to properly identifying the heterogeneous population of patients with HFrEF (10). In this study, we developed and externally validated a state-of-the-art deep-learning language model that identifies HFrEF through contextual analysis of clinical notes. Our model can effectively process long notes without being limited to pre- defined terms. Our approach suggests a computable phenotype of HFrEF from unstructured EHR documents, which is an essential step to automate quality-of-care measurement and improvement strategies in heart failure.

## METHODS

The study was reviewed by the Yale Institutional Review Board, which approved the study protocol and waived the need for informed consent, as the study represents a secondary analysis of existing data.

### Data Source for Model Development

The model was developed using the EHR data of the Yale New Haven Hospital (YNHH) from 2015 through 2019. YNHH is a large academic hospital that serves a diverse population with a significant proportion of underrepresented minorities. The EHR vendor at YNHH is Epic^®^, and we obtained and curated an extract spanning structured and unstructured data fields for the study population. The structured fields included patient demographics, diagnosis and procedure codes, medications, and healthcare encounters. The unstructured data included notes, such as history and physical exam (H&P), progress reports, procedure reports, and discharge summaries. In addition, the information was merged with the data on echocardiography parameters, which were available from our structured echocardiography reporting system (Lumedx^®^, Oakland, CA).

### Study Population

The study population included all individuals hospitalized with a principal or secondary diagnosis of heart failure with at least one echocardiogram during or before the index hospitalization between 2015 and 2019. Heart failure hospitalizations were defined using international classification of diseases (ICD) diagnosis codes (**Table S1**). To maximize learning with the broadest set of available data, hospitalizations as opposed to patients were chosen as the unit of analysis.

Discharge summaries from hospitalizations with heart failure were obtained and used for model development and validation. These notes encapsulate the summary of patient’s hospitalization, including the presenting symptoms, hospital course, diagnostic studies, and therapeutic interventions, and contain rich evidence to support a clinical diagnosis.

### Study Outcomes and Covariates

Diagnosis of HFrEF was the primary outcome of the study. This diagnosis was ascertained based on left ventricular ejection fraction (LVEF) < 40% on antecedent echocardiography before or during the index hospitalization.

The proportional use of guideline-directed medical therapies among model-based reclassified patients with HFrEF was the secondary outcome of the study. These therapies included beta blockers, angiotensin receptor-neprilysin inhibitor (ARNI), angiotensin-converting enzyme inhibitors (ACEI), angiotensin receptor blockers (ARB), sodium-glucose cotransporter-2 inhibitors (SGLT2i), and mineralocorticoid receptor blockers (MRA) based on recommendations from the 2022 AHA/ACC/HFSA Guideline for the Management of Heart Failure (7). **Table S2** represents the medications in each drug class and the adjudication strategy to determine their use during hospitalization or prescription at the time of discharge. The proportional use of each drug class was calculated among individuals who were correctly reclassified as HFrEF based on model predictions compared with chart diagnosis codes with LVEF on antecedent echocardiography as the gold standard for the diagnosis of HFrEF. Demographic characteristics of the study population, including age, sex, and race were collected from the database. Categories of race included non-Hispanic White, non-Hispanic Black, Hispanic, and individuals of other races.

### Data Processing and Model Development

We developed a deep learning-based language model built upon a pretrained clinical longformer to identify individuals with HFrEF (11). This model takes advantage of an attention mechanism known as sliding window attention, which enables the algorithm to process text sequences by using multiple stacked layers of small window attentions surrounding each token (12). The longformer provides contextual insights through processing long-term dependencies in the entire clinical note, and has previously achieved state-of-the art results on a range of clinical tasks, including named-entity recognition, question answering, natural language inference, and document classification tasks. Such attention-based models enable understanding of the context in which each term is used in the text, including the presence or absence of a condition (negation detection), historical feature, or the reference to the presence of a condition in some other individual (such as family, instead of the patient).

We randomly split notes into train (70%) and test (30%) sets after stratification at the patient level to avoid sharing data from the same patient between two sets. Punctuations, special characters, and stop words were removed from the corpus of each note and the remaining string input were tokenized and converted to tensors. A maximum input sequence length of 4,096 words was set based on the maximum input length allowed for the base model (11), and a truncation function was applied to tensors longer than this limit (**Figure 1)**.

**Figure 1.**
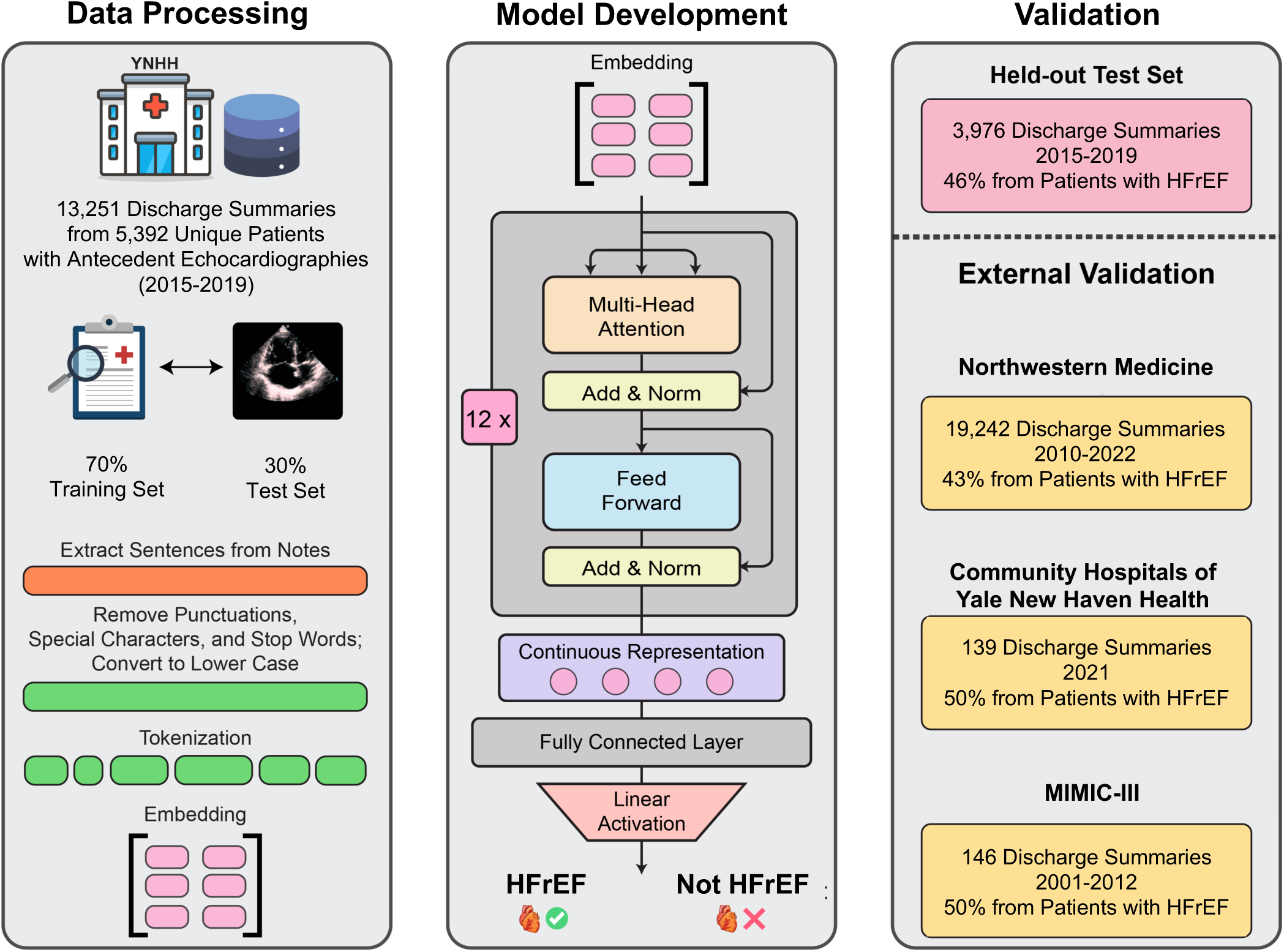
Model Development. Hospitalizations for heart failure at Yale New Haven Hospital with an antecedent echocardiography were included for model development. The schematic represents data processing, model development, and evaluation of model performance on validation sets, including Northwestern Medicine, community hospitals of Yale New Haven health system, and MIMIC-III database. Abbreviations: YNHH, Yale New Haven Hospital; HFrEF, heart failure with reduced ejection fraction; MIMIC, Medical Information Mart for Intensive Care.

The model weights were initialized with pre-trained values and fine-tuned on input embeddings using a backpropagation gradient descent algorithm with a cross entropy loss function. A single linear layer perceptron was applied to the output of the longformer for binary classification. This layer receives the last hidden state of the longformer and predicts the probability of HFrEF using a Softmax activation function. Adam optimizer was used for stochastic optimization without any drop-out. The model was trained with a learning rate of 1e-7 for 30 epochs, with early stopping rules set based on lack of improvement of validation loss for 3 consecutive epochs, upon which the preceding epoch was chosen as the final study model.

### Validation

The model’s performance in detecting individuals with HFrEF was internally assessed on the held-out test set. The diagnosis of HFrEF was ascertained based on LVEF < 40% on antecedent echocardiography, as described above. We also evaluated the association of model-predicted probabilities of HFrEF with echocardiography-based measurements of LVEF to assess the plausibility of model predictions. In sensitivity analyses, the model was validated on clinical subgroups of cardiovascular comorbidities, including ischemic heart disease, atrial fibrillation/flutter, and right heart failure, which were defined based on corresponding disease diagnosis codes (**Table S1**). We also assessed the performance of the model on discharge summaries of variable lengths, as determined by the quintiles of word counts of these notes.

Furthermore, we evaluated the model’s performance after blanking out echocardiography report sections from discharge summaries. This was designed to evaluate the broader generalizability of our model to practice locations where automated inclusion of echocardiographic reports is not the norm. For this analysis, we utilized a custom dictionary to identify structured section headers within the body of the notes. Leveraging an entity recognition module in MedSpaCy library, we set up a rule-based pipeline to identify echocardiography section header terms (13). To enhance the performance of this approach, we fine-tuned a BioClinicalBERT algorithm, a publicly available language model specialized for biomedical text analysis (14), by training the original model on a labeled dataset where echocardiography sections were tagged in the discharge summaries. The rule-based entity recognition module served as a preliminary filter, after which fine-tuned BioClinicalBERT model assessed the likelihood of a flagged section containing echocardiography data.

In addition to the echocardiogram-based label, a sample of 130 discharge summaries was randomly selected from the held-out test set and manually reviewed by a cardiologist for a diagnosis of HFrEF. The phenotype of heart failure was adjudicated in a blind fashion to model predictions based on patient’s signs and symptoms, lab values, and LV systolic function in the body of discharge summaries. A diagnosis of HFrEF was ultimately made in 58 notes (45%) based on clinical judgement of the adjudicating cardiologist.

### Data Source: External Validation

To assess the generalizability of the model, we validated our algorithm in three external sets, including: 1) Northwestern Medicine health system; 2) community hospitals of Yale New Haven health system in the states of Connecticut and Rhode Island; and 3) hospitalizations requiring intensive care from Medical Information Mart for Intensive Care III (MIMIC-III) database.

Hospitalizations with heart failure at Northwestern Medicine, a large integrated health system in Illinois, USA, were identified based on a principal diagnosis of heart failure between January 2010 to December 2022. All individuals with a preceding echocardiography within 6 months of the discharge were included in this analysis. LVEF < 40% on the most recent echocardiography was used to define the HFrEF label. This sample included 19,242 notes from 11,513 unique individuals, including 8,277 (43%) notes from patients with LVEF < 40%. These analyses were performed in a fully federated fashion by the collaborators, without access to the data by the model development team at Yale.

In addition, we identified hospitalizations with heart failure at satellite community hospitals of the Yale New Haven health system between January to December 2021. These hospitals include Bridgeport Hospital, Lawrence & Memorial Hospital, and Greenwich Hospital in Connecticut, and Westerly Hospital in Rhode Island. Hospitalizations with a principal or secondary diagnosis of heart failure were identified from diagnosis codes, consistent with the prior analysis (**Table S1)**. This sample included 139 discharge summaries manually reviewed by a cardiologist and a diagnosis of HFrEF was adjudicated in 70 (50.3%) notes. The manual review was blind to model predictions and patient-level data beyond the note.

Finally, we validated the model on discharge summaries from MIMIC-III, a database of de-identified medical records from admissions that required critical care during their hospitalization at Beth Israel Deaconess Medical Center between 2001 to 2012 (15). MIMIC-III database houses unstructured notes and structured data fields, such as patient demographics, laboratory tests, in-hospital procedures, and prescription medications. A random sample of 146 discharge summaries was selected from the original dataset based on a principal discharge diagnosis of heart failure, and manually reviewed by a cardiologist, which identified a diagnosis of HFrEF in 74 (50.6%) of notes.

### Comparison with Chart Diagnosis

We assessed the model’s performance in reclassifying HFrEF phenotype from discharge summaries compared with the chart diagnosis codes. An LVEF < 40% on antecedent echocardiography was considered the true label for HFrEF. Chart diagnosis codes corresponding with systolic heart failure were used in this analysis and are presented in **Table S1**. The proportional use of guideline-directed therapies among reclassified individuals was assessed as a secondary outcome, as described in section “study outcomes and covariates” above.

### Explainability Analyses

We performed an explainability analysis using Local Interpretable Model-agnostic Explanations (LIME) to identify predictive keywords for HFrEF from discharge summaries. This analysis estimates the contribution of local regions of the input text by perturbating input instances while observing the corresponding changes in model predictions (16). LIME-generated estimates capture local dependencies and provide insights into the feature importance for model predictions. We applied LIME analysis to the top 100 most confident model predictions of HFrEF and extracted the top 20 most predictive keywords for HFrEF based on their corresponding weights. We also demonstrate representative examples of this analysis on real-world discharge summaries that represents the relative importance of each token in the final model prediction.

### Statistical Analysis

Categorical variables are presented in frequency and percentage. Continuous variables are presented in mean and standard deviation or median and interquartile range, as appropriate. Area under receiver operating characteristic (AUROC) was used to assess model discrimination. Sensitivity, specificity, positive predictive value (PPV), negative predictive value (NPV), and area under precision-recall curve (AUPRC) were calculated to assess the model performance. The cut-off probability for binary prediction of HFrEF was set at 0.5 and was consistent across all internal and external validations.

To compare the performance of the model with chart diagnosis classification of HFrEF, the AUROCs of the two methods were compared. The AUROC and corresponding 95% confidence intervals for this analysis were obtained from a logistic regression model with the binary prediction of HFrEF as the outcome. A univariate linear regression was used to assess the association between LVEF values as outcome and model-predicted probabilities of HFrEF as predictor. Model-predicted probabilities of HFrEF were transformed on a natural logarithmic scale for this analysis.

Net reclassification improvement (NRI) was used to assess model-based reclassification of HFrEF compared with the chart diagnosis codes. NRI is a statistical method to assess the improvement in disease classification by risk prediction models and quantifies correct reclassification of individuals into more appropriate risk or disease categories. Proportional use of guideline-directed therapies was aggregated for medications in the same drug class among patients who were correctly reclassified to HFrEF after applying model predictions to chart diagnosis codes.

All statistical tests were 2-sided with a level of significance of 0.05. Computational and statistical analyses were performed in Python 3.8. Data processing was performed using Pandas (1.5.3), Numpy (1.23.5), SciPy (1.10.1), and Scikit-learn (1.2.2). Model development was performed using Pytorch (1.4.0). The pretrained clinical longformer model was imported from the Hugging Face transformers library. LIME library (0.2.0) was used for explainability analysis. SpaCy (3.5) and MedSpaCy (0.2.0) were used for fine-tuning the BioClinicalBERT model for identification of echocardiography sections in discharge summaries. (**Table S3**).

## RESULTS

### Study Population

There were 13,251 hospitalizations for heart failure from 5,392 unique patients at YNHH between 2015 to 2019. Discharge summaries of these hospitalizations represented training (n=9,275, 70% of notes) and held-out test sets (n=3,976, 30% of notes). The mean age of the study population was 73 ± 14 years, and 48% were female. Overall, 3,721 individuals were non-Hispanic White (69.0%), 1,118 were non-Hispanic Black (20.7%), 386 were Hispanic (7.1%), and 167 self-identified as other races (3.0%). A total of 2,487 patients (46.1%) had LVEF of less than 40%, 736 (13.6%) had LVEF between 40% to 50%, and 2,169 (40.2%) had LVEF of 50% or greater. Overall, 78.3% were treated with a beta-blocker, 9.4% with sacubitril-valsartan, 28.9% with an ACEI, 19.3% with an ARB, 2.6% with an SGLT-2i, and 26.3% with an MRA. The demographic and clinical characteristics of the study population are presented in **Table 1**.

**Table 1.**
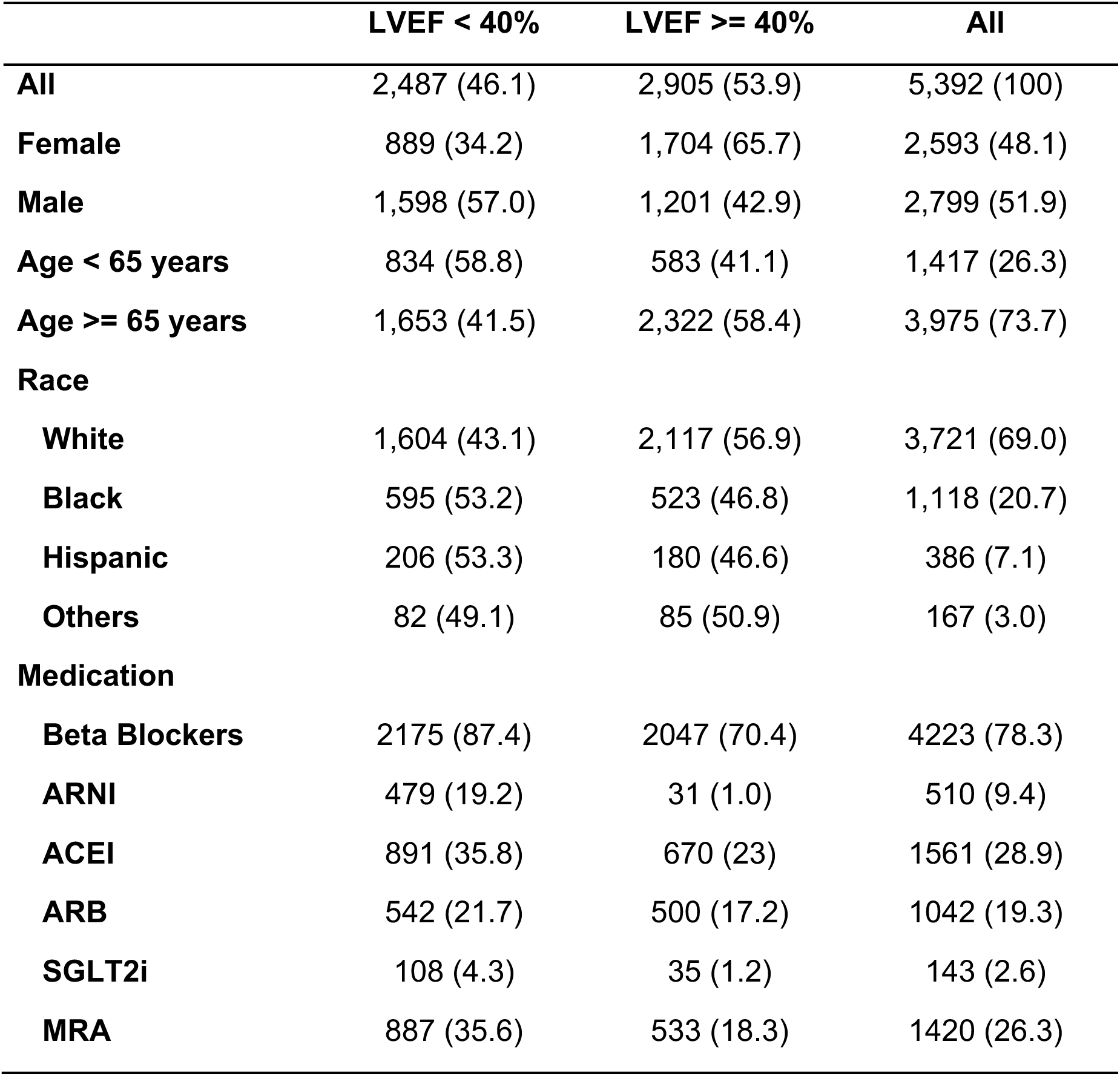
Characteristics of the Study Population. Values are presented as number (percentage). Medications include guideline-directed medical therapies during or at the time of discharge from the index hospitalization at Yale New Haven Hospital. Patients receiving sacubitril-valsartan were included in both ARNI and ARB groups. Abbreviation: LVEF, left ventricular ejection fraction; ARNI, angiotensin receptor-neprilysin inhibitor; ACEI, angiotensin converting enzyme inhibitor; ARB, angiotensin receptor blocker; SGLT2i, sodium glucose cotransporter 2 inhibitor; MRA, mineralocorticoid receptor antagonist.

### Detection of HFrEF from Clinical Notes

The model demonstrated an AUROC of 0.97 and AUPRC of 0.97 in detecting individuals with HFrEF on the held-out test set based on a preceding echocardiogram with LVEF < 40% (**Figure 2**). The sensitivity and specificity of the model were 0.89 and 0.94, with PPV and NPV of 0.93 and 0.90, respectively. Model performance was consistent across demographic subgroups of age, sex, and race (**Table 2)**.

**Figure 2.**
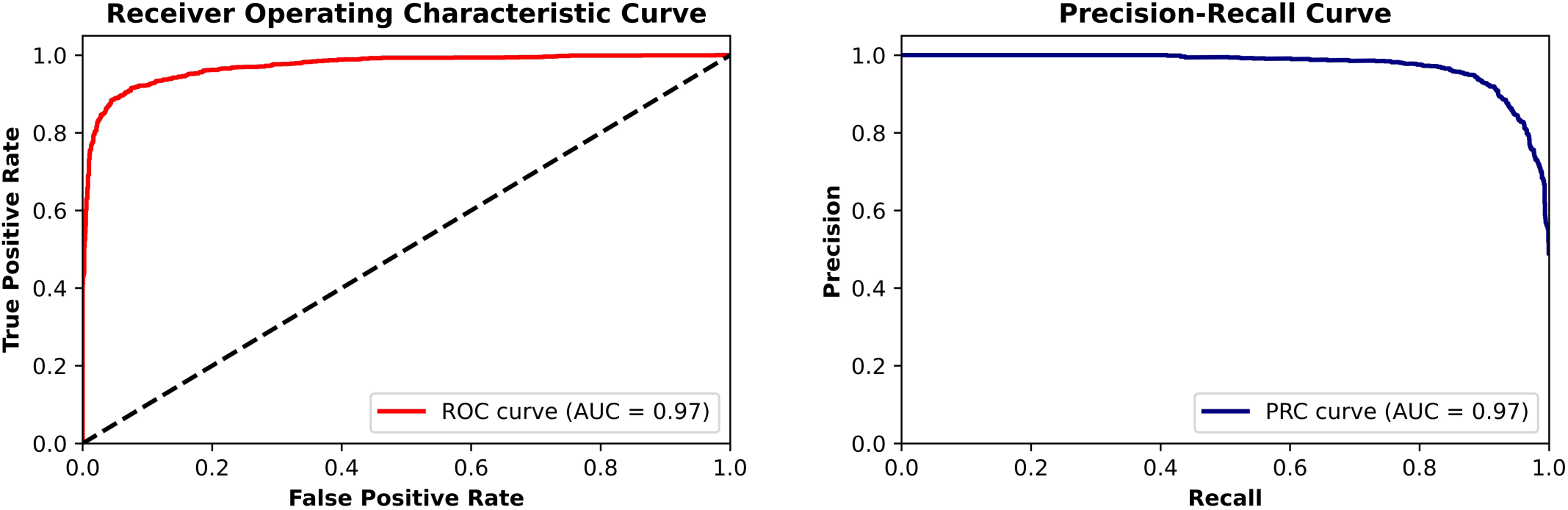
Model Performance. The graphs represent receiver operating characteristic (left) and precision-recall (right) curves in identifying HFrEF on clinical notes from the held-out test set at Yale New Haven Hospital. Abbreviations: ROC, receiver operating characteristic curve; PRC, precision recall curve; AUC, area under curve.

**Table 2.** Model Performance Across Demographic Subgroups. Data represent performance metrics for detecting HFrEF across subgroups of age, sex, and race on the held-out test set at Yale New Haven Hospital. Abbreviations: AUROC, area under receiver operating characteristic curve; AUPRC, area under precision recall curve; PPV, positive predictive value; NPV, negative predictive value.

|  | **AUROC** | **AUPRC** | **Sensitivity** | **Specificity** | **PPV** | **NPV** |
| --- | --- | --- | --- | --- | --- | --- |
| **All** | 0.971 | 0.973 | 0.896 | 0.941 | 0.934 | 0.906 |
| **Female** | 0.972 | 0.956 | 0.877 | 0.962 | 0.920 | 0.940 |
| **Male** | 0.970 | 0.980 | 0.910 | 0.908 | 0.935 | 0.874 |
| **Age < 65** | 0.979 | 0.986 | 0.946 | 0.900 | 0.937 | 0.914 |
| **Age >= 65** | 0.970 | 0.967 | 0.880 | 0.948 | 0.927 | 0.912 |
| **White** | 0.968 | 0.966 | 0.883 | 0.943 | 0.925 | 0.910 |
| **Black** | 0.987 | 0.990 | 0.945 | 0.930 | 0.939 | 0.936 |
| **Hispanic** | 0.973 | 0.978 | 0.928 | 0.934 | 0.859 | 0.914 |
| **Others** | 0.984 | 0.984 | 0.909 | 0.963 | 0.821 | 0.913 |

In the confirmatory analysis of 130 expert-adjudicated notes from the held-out test set, the model had an AUROC of 0.99 and AUPRC of 0.99 in detecting HFrEF, with sensitivity and specificity of 0.91 and 0.97, and PPV and NPV of 0.96 and 0.93, respectively.

### External Validation

There were 19,242 notes from 11,513 unique individuals in the validation set from Northwestern Medicine, including 5,386 (46.7%) women. Overall, 7,460 individuals were non-Hispanic White (64.7%), 2,466 were non-Hispanic Black (21.4%), 663 were Hispanic (5.7%), and 954 self-identified as other races (8.2%). In this sample, 8,277 (43%) notes were for patients with an LVEF < 40% on an antecedent echocardiography. In this validation set, the model demonstrated an AUROC of 0.94 and AUPRC of 0.91 in detecting HFrEF, with sensitivity and specificity of 0.83 and 0.89, and PPV and NPV of 0.86 and 0.87, respectively (**Figure 3**).

**Figure 3.**
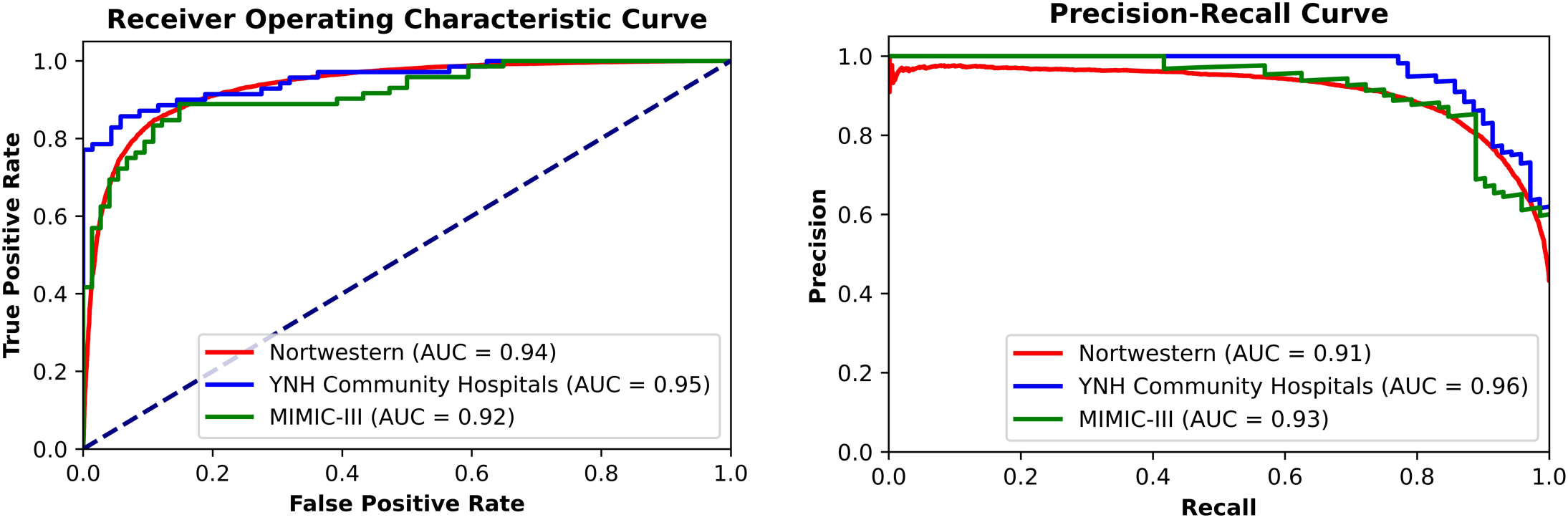
External Validation. Performance of the model in identifying HFrEF was evaluated on discharge summaries from three external sets, including 1) Northwestern Medicine, 2) Community hospital of Yale New Haven health system, and 3) MIMIC-III. The figure represents receiver operating characteristic (left) and precision-recall (right) curves for these analyses. Abbreviations: YNH, Yale New Haven; MIMIC, Medical Information Mart for Intensive Care III; AUC, area under curve.

We further validated the model on manually reviewed discharge summaries from community hospitals of Yale New Haven Health System (n=139) and MIMIC-III database (n=146). Individuals in both datasets were predominantly male (61.9% and 65.7%), non-Hispanic White (77.6% and 71.2%), and older than 65 years of age (79.2% and 74.6%, respectively). A diagnosis of HFrEF was adjudicated in half of the notes from each validation set (50%). The model demonstrated an AUROC of 0.95 and AUPRC of 0.96 in detecting HFrEF from clinical notes of Yale community hospitals. The sensitivity and specificity of the model in identifying HFrEF were 0.77 and 0.98, with PPV and NPV of 0.98 and 0.80, respectively. On the MIMIC-III dataset, the model had an AUROC of 0.91 and AUPRC of 0.92 in detecting HFrEF, with sensitivity and specificity of 0.77 and 0.90, and PPV and NPV of 0.88 and 0.80, respectively. These PPV and NPV were at the a priori probability threshold of 0.5. Baseline characteristics of individuals in each dataset and model performance on notes from three validation sets are presented in **Table S4** and **Figure 4**.

**Figure 4.**
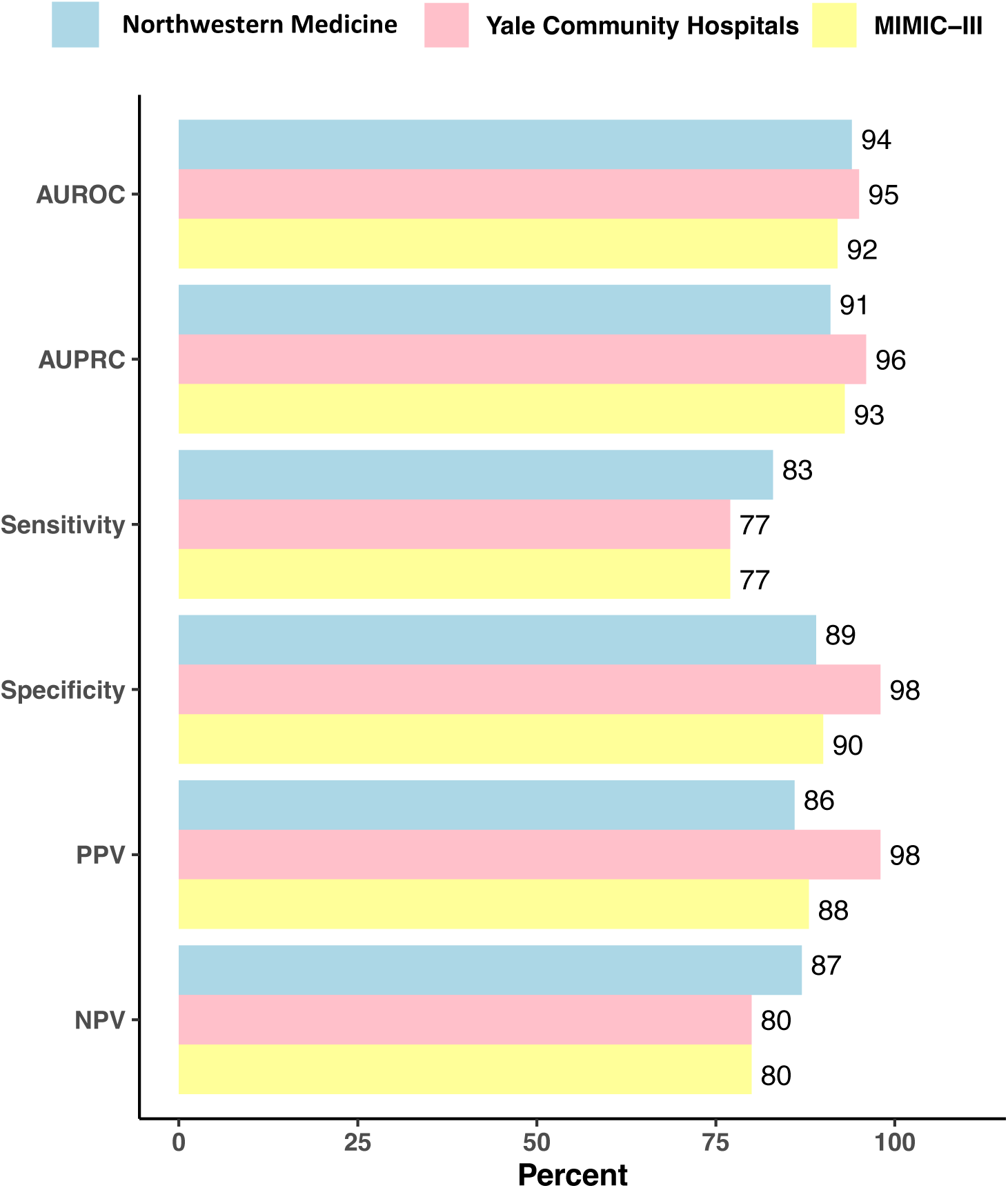
External Validation. The bar graph represents model performance in detecting HFrEF from discharge summaries of hospitalizations with heart failure at Northwestern Medicine (blue), community hospitals of Yale New Haven health (pink) and MIMIC-III (yellow). Abbreviations: AUROC, area under receiving operating characteristics; AUPRC, area under precision recall curve; PPV, positive predictive value; NPV, negative predictive value; MIMIC, Medical Information Mart for Intensive Care III.

### Model Performance across Cardiovascular Comorbidities, Notes of Varying Lengths, and Discharge Summaries Without Echocardiography Reports

In sensitivity analyses, the performance of the model in detecting HFrEF from discharge summaries in the held-out test set remained consistent among patients with or without ischemic heart disease (AUROC 0.96 vs 0.97, AUPRC 0.98 vs 0.97, respectively), atrial fibrillation/flutter (AUROC 0.97 and AUPRC 0.98 in both subgroups), and right heart failure (AUROC 0.99 vs 0.97, AUPRC 0.97 vs 0.97, respectively). Receiver operating characteristics and precision-recall curves for these analyses are presented in **Figures S3-S5**.

In further sensitivity analyses, we evaluated the model’s performance on clinical notes of varying lengths. The median word count of discharge summaries in the held-out test set was 3,192 [IQR 1,604-4,922], and 58% of notes had 4,096 words or fewer, the truncation threshold for longer notes in our data processing pipeline. Among notes shorter than this limit, 995 (43%) belonged to individuals with HFrEF, compared with 925 (55.4%) notes longer than this cut point. The performance of the model in detecting HFrEF was consistent across the quintiles of word count, with AUROC ranging between 0.96-0.98 and AUPRC between 0.94-0.99 (**Table S5**).

After blanking out echocardiography report sections from the discharge summaries, the model had similar performance in identifying HFrEF with an AUROC of 0.94 and AUPRC of 0.94, sensitivity of 0.86, specificity of 0.89, PPV of 0.88, and NPV of 0.87.

### Disease Reclassification and Use of Guideline-directed Therapies

The model correctly identified individuals with low LVEF in 43% and normal LVEF in 49% of notes in the held-out test set, compared with 18% and 44% with corresponding chart diagnosis codes, respectively (**Figure 5A and Table S6)**. Model-based predictions of HFrEF corresponded to an overall NRI of 60.2 ± 1.9% compared with the chart diagnosis codes (p-value < 0.001), and an increase in AUROC from 0.61 [95% CI: 0.60- 0.63] to 0.91 [95% CI 0.90-0.92] with binary predictions (**Table S7**).

**Figure 5.**
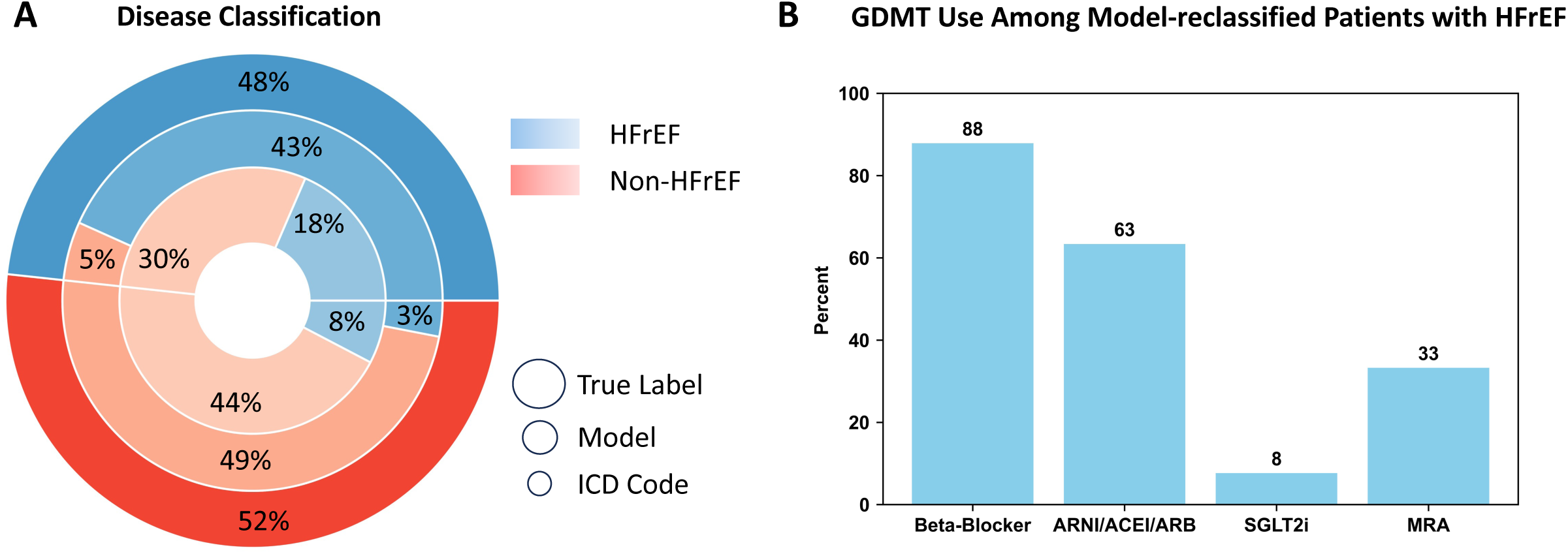
Disease Classification and Use of Guideline Directed Therapies. (A) Nested pie chart demonstrates disease classification based on echocardiography measurements of LVEF (true label, outer circle), model predictions of disease phenotype (middle circle), and chart-documented diagnosis codes (inner circle). Blue represents a diagnosis of HFrEF and red represents other phenotypes of heart failure; (B) Use of guideline-directed therapies among individuals with LVEF < 40% who were correctly reclassified from a chart-diagnosis of non-HFrEF to a model-based diagnosis of HFrEF. Abbreviations: ARNI, angiotensin receptor-neprilysin inhibitor; ARB, angiotensin receptor blocker; ACEI, angiotensin converting enzyme inhibitor; SGLT2i, sodium glucose cotransporter 2 inhibitor; MRA, mineralocorticoid receptor antagonist.

The proportional use of guideline-directed therapies among individuals who were correctly reclassified to HFrEF after applying the model is presented in **Figure 5B**. In this group of individuals, 88% were treated with a beta-blocker, 63% with ARNI, ACEI, or ARB, 8% with an SGLT2i, and 33% with an MRA.

### Explainability Analysis

In an analysis of textual terms associated with HFrEF, we identified the top 20 keywords within a clinical note predictive of a HFrEF phenotype. The top 3 predictive words for the presence of HFrEF were “severe”, “globally”, and “dobutamine”. This exploratory analysis identified clinically relevant terms, such as “HFrEF”, “ventricular assist device (VAD)”, “entresto”, and “inotropic” from the clinical note that predict a HFrEF phenotype (**Figure 6**). A representative example of this analysis on a discharge summary from an individual with HFrEF is shown in **Figure 6B**.

**Figure 6.**
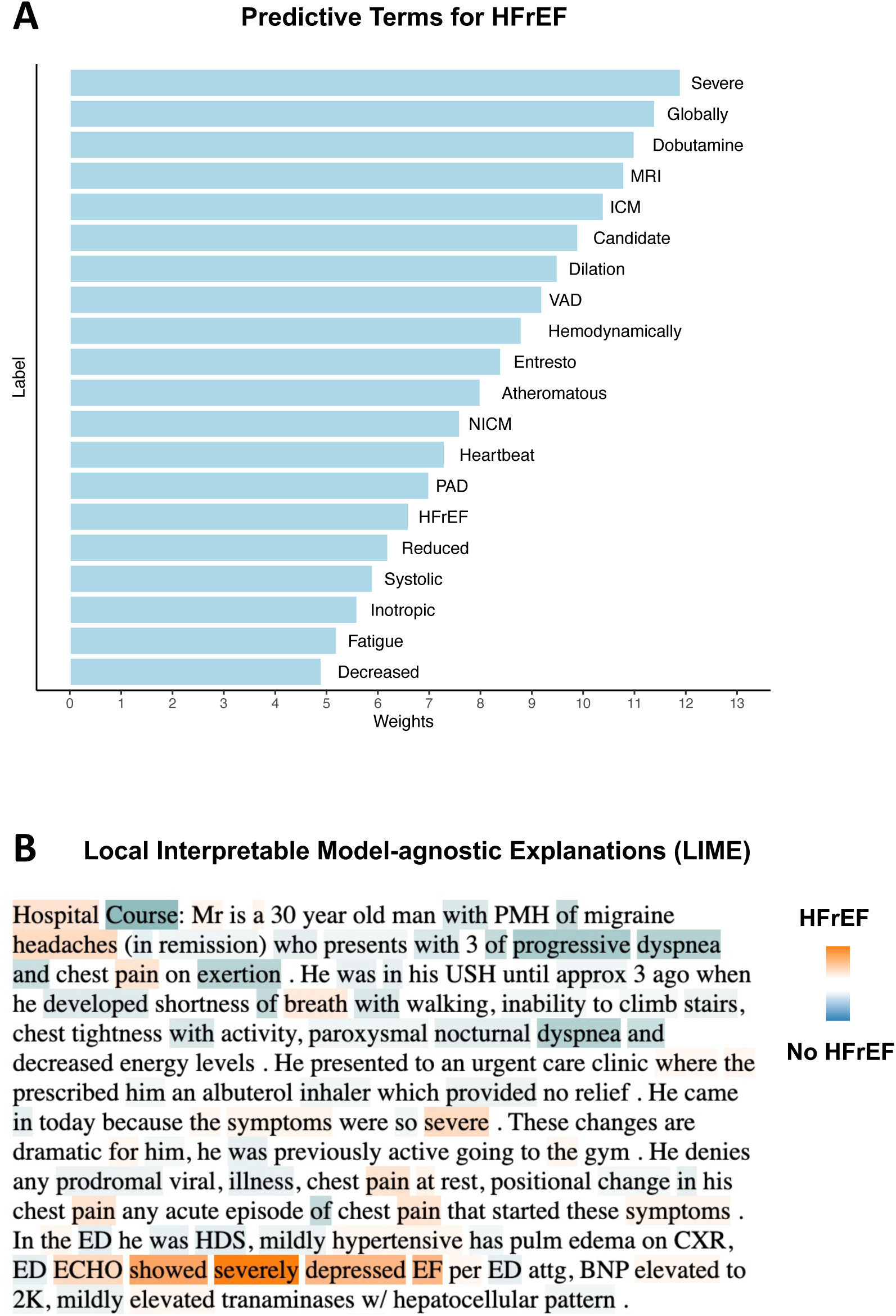
Predictive Keywords for Heart failure Phenotypes. Figure represents Local Interpretable Model-agnostic Explanations (LIME) for model predictions of HFrEF from discharge summaries. **(A)** The most predictive keywords for HFrEF with corresponding coefficients based on the LIME analysis of the top 100 notes with the most confident model predictions. **(B)** A real-world example of LIME analysis on a discharge summary representing local dependencies. The color intensity of each highlighted keyword represents its contributions to the model-predicted probability of a HFrEF (orange) vs a non-HFrEF phenotype (blue). The deidentified discharge summary describes the hospitalization of an individual after initial presentation with heart failure and left ventricular ejection fraction of 30% on echocardiography. The model predicted a 99% probability for a HFrEF from this discharge summary. Abbreviations: MRI, magnetic resonance imaging; ICM, ischemic cardiomyopathy; VAD, ventricular assist device; NICM, non-ischemic cardiomyopathy; PAD, peripheral arterial disease; HFrEF, heart failure with reduced ejection fraction; PMH, past medical history; USH, usual state of health; ED, emergency department; HDS, hemodynamically stable; CXR, chest x-ray; ECHO, echocardiography; EF, ejection fraction; BNP, B-natriuretic peptide.

## DISCUSSION

We developed and externally validated a deep learning-based natural language processing model that automatically detects HFrEF from clinical notes in the EHR. The algorithm demonstrated high discrimination in identifying HFrEF across demographic subgroups of age, sex, race, cardiovascular comorbidities, and discharge summaries of varying lengths. It also showed a robust performance across multiple regionally and temporally distinct academic and community clinical settings. This strategy improved the classification of patients with HFrEF compared with the chart diagnosis codes and identified discernible gaps in guideline-directed therapies in practice. Moreover, our language-based approach is explainable, as it detects clinically relevant terms through contextual analysis of clinical notes. In combination with diagnosis codes and echocardiography, this algorithm can systematically identify patients with HFrEF who would benefit from guideline-directed treatments.

Accurate Identification of heart failure subtypes is essential to effectively implement guideline-recommended therapies in practice, as evidence-based treatments can significantly vary across phenotypes of heart failure (17). The current workflow to assess the quality of care in heart failure registries relies on abstractors to manually collect information on patients with heart failure at participating hospitals, a resource-intensive strategy with challenges in scaling at the national level and improving the quality of care in real-time (18). Prior population-based studies have relied on administrative claims data and diagnosis codes to identify individuals with heart failure but only achieved limited accuracy with sensitivity ranging between 68-72% and specificity between 63-68% in identifying heart failure subgroups such as HFrEF (19). These approaches do not use unstructured EHR data to discern heart failure phenotypes, as leveraging this resource has been difficult to implement in practice (20). Our model, however, is a powerful tool to exploit the previously underused data in clinical notes to improve the quality of care in this population.

Previous applications of NLP in heart failure were limited to rule-based models to extract LVEF from echocardiography reports (21), search for pre-specified terms in notes (22), or annotate medical conditions from clinical notes with partitioned format (23). The real-world applications of these methods remain limited due to variations of EHR format across institutions. In addition, the dynamic changes of echocardiography parameters such as LVEF over time pose a temporal challenge to ascertaining heart failure subtypes from cross-sectional measurements. Our method overcomes these challenges through contextual analysis of the entire note without restriction to pre-specified keywords. The model learned to detect the lexical predictors of HFrEF, which were consistent with the clinical characteristics of this condition, suggesting the explainability of the model predictions. Moreover, our approach does not require partitioning or segmentation of the input text and is hence applicable independent of the EHR format.

The fully automated nature of our approach is not its only advantage over the current methods of quality assessments. In identifying individuals with HFrEF, our model outperformed the diagnosis codes by a large margin. It demonstrates consistent performance across geographically distant settings with no requirement for reformatting the model before implementation. Our approach could not only be an adjunct to the structured methods of identifying individuals with HFrEF but also applicable to settings where echocardiography-based measurements of LVEF are not systematically collected. On a broader scale, our findings suggest the immense potential of artificial intelligence in understanding human language and its applications in health care to promote the quality of care in clinical practice.

Our study has some limitations that merit consideration. First, the performance of our model in detecting HFrEF depends on the quality of chart documentation. Poorly documented hospital courses or patient conditions would predictably lead to imperfect model performances. This limitation is not exclusive to our approach, as manual chart abstractions encounter similar challenges in identifying patient subgroups. Second, our model is not prospectively validated to monitor the quality of care. However, the generalizability of our approach is supported by validation in multiple sites, and the gaps in the use of guideline-directed therapies provide an actionable target to improve the quality of care compared with the chart diagnosis codes. Third, the proportional use of guideline-directed therapies as the secondary outcome of the study does not consider eligibility for treatment, as renal dysfunction, electrolyte abnormalities, or hypotension could potentially impact the treatment decisions for the corresponding medications. The explainability analysis of our model predictions also suggest clinical features of individuals who may not be candidate for these therapies, as this analysis was limited to 100 individuals with the most confident model predictions of HFrEF with overrepresentation of patients with advanced heart failure. However, the observed gaps in the use of guideline-directed therapies in our study population was consistent with previous reports of similar national trends (3,4). Further analysis of treatment eligibilities might be necessary to distinguish individuals with advanced heart failure from those who would benefit from the initiation of guideline-directed therapies.

## CONCLUSION

We developed an automated algorithm that identifies patients with HFrEF through an explainable, deep learning-based natural language processing of clinical notes in the EHR. Our approach improved the appropriate classification of patients with heart failure compared with the chart diagnosis codes and suggested a novel computable phenotype of HFrEF from EHR that could be used for quality assessment and improvement.

## Data Availability

All data produced in the present work are contained in the manuscript.

## Funding

This study was supported by research funding awarded to Dr. Khera by the Yale School of Medicine and grant support from the National Heart, Lung, and Blood Institute of the National Institutes of Health under the award K23HL153775. The funders had no role in the design and conduct of the study; collection, management, analysis, and interpretation of the data; preparation, review, or approval of the manuscript; and decision to submit the manuscript for publication.

## Disclosures

Dr. Tariq Ahmad is a consultant for Sanofi-Aventis, Amgen, and Cytokinetics. He has research funding from Boehringer Ingelheim, AstraZeneca, Cytokinetics, and Relypsa. Dr. Nadkarni reports consultancy agreements with AstraZeneca, BioVie, GLG Consulting, Pensieve Health, Reata, Renalytix, Siemens Healthineers and Variant Bio; has received research funding from Goldfinch Bio, and Renalytix; has received honoraria from AstraZeneca, BioVie, Lexicon, Daiichi Sankyo, Meanrini Health, and Reata; has patents or royalties with Renalytix; owns equity and stock options in Pensieve Health and Renalytix as a scientific cofounder; owns equity in Verici Dx; has received financial compensation as a scientific board member and advisor to Renalytix; has served on the advisory board of Neurona Health; and has served in an advisory or leadership role for Pensieve Health and Renalytix. Dr. Faraz Ahmad reports grants from the Agency for Healthcare Research and Quality, grants from the National Institutes of Health/National Heart, Lung, and Blood Institute, grants from the American Heart Association, personal fees from Teladoc, personal fees from Livongo, personal fees from Pfizer, outside the submitted work. Dr. Krumholz works under contract with the Centers for Medicare & Medicaid Services to support quality measurement programs, was a recipient of a research grant from Johnson & Johnson, through Yale University, to support clinical trial data sharing; was a recipient of a research agreement, through Yale University, from the Shenzhen Center for Health Information for work to advance intelligent disease prevention and health promotion; collaborates with the National Center for Cardiovascular Diseases in Beijing; receives payment from the Arnold & Porter Law Firm for work related to the Sanofi clopidogrel litigation, from the Martin Baughman Law Firm for work related to the Cook Celect IVC filter litigation, and from the Siegfried and Jensen Law Firm for work related to Vioxx litigation; chairs a Cardiac Scientific Advisory Board for UnitedHealth; was a member of the IBM Watson Health Life Sciences Board; is a member of the Advisory Board for Element Science, the Advisory Board for Facebook, and the Physician Advisory Board for Aetna; and is the co-founder of Hugo Health, a personal health information platform, and co-founder of Refactor Health, a healthcare AI-augmented data management company. Dr. Khera is an Associate Editor of JAMA. He receives support from the National Heart, Lung, and Blood Institute of the National Institutes of Health (under award K23HL153775) and the Doris Duke Charitable Foundation (under award, 2022060). He also receives research support, through Yale, from Bristol-Myers Squibb and Novo Nordisk. He is a coinventor of U.S. Provisional Patent Applications 63/177,117, 63/428,569, 63/346,610, 63/484,426, and 3/508,315. He is a co-founder of Evidence2Health, a precision health platform. The remaining authors have no competing interests to disclose.

