## Supplemental File for "Automated Identification of Heart Failure with Reduced Ejection Fraction using Deep Learning-based Natural Language Processing"

**Table S1. International Classification of Diseases Diagnosis Codes for Identification of Patient Groups.** Abbreviation: ICD, international classification of disease.

| **Diagnosis** | **ICD-9** | **ICD-10** |
|---|---|---|
| **Heart Failure** | 402.x, 404.x, and 428.x | I11.0, I13.0, I30.2, and I50.x |
| **Systolic Heart Failure** | 428.2 and 428.4 | I50.2 and I50.4 |
| **Ischemic Heart Disease** | 414.x | I25.x |
| **Atrial Fibrillation/Flutter** | 427.3 | I48.x |
| **Right Heart Failure** | N/A | I50.81 |

**Table S2. Guideline Directed Medical Therapies for HFrEF.** Proportional use of each drug class was determined by searching the names of medications in the corresponding class among all administered drugs during the index hospitalization or prescribed medications at the time of discharge. Abbreviations: ARNI, angiotensin receptor-neprilysin inhibitor; ACEI, angiotensin converting enzyme inhibitor; ARB, angiotensin receptor blocker; MRA, mineralocorticoid receptor antagonist; SGLT2i, sodium glucose cotransporter 2 inhibitor.

| Beta Blockers | ARNI | ACEI | ARB | MRA | SGLT2i |
|---|---|---|---|---|---|
| Bisoprolol | Sacubitril-Valsartan | Captopril | Candesartan | Spironolactone | Dapagliflozin |
| Carvedilol | | Enalapril | Losartan | Eplerenone | Empagliflozin |
| Metoprolol Succinate | | Fosinopril | Valsartan | | |
| | | Lisinopril | | | |
| | | Quinapril | | | |
| | | Ramipril | | | |
| | | Trandolapril | | | |
| | | Perindopril | | | |

**Table S3. Analytic Packages and Languages Used for Model Development and Statistical Analysis**

| Programming Language/Package | Version |
|---|---|
| Python | 3.8 |
| Pandas | 1.5.3 |
| Numpy | 1.23.5 |
| SciPy | 1.10.1 |
| Scikit-learn | 1.2.2 |
| LIME | 0.2.0 |
| Pytorch | 1.4.0 |
| SpaCy | 3.5 |
| MedSpaCy | 0.2.0 |

**Table S4. Characteristics of Patient Populations in Validation Sets.** Values are presented as number (percentage). Abbreviations: YNH, Yale New Haven health system; MIMIC, Medical Information Mart for Intensive Care; LVEF, left ventricular ejection fraction. *14 patients with unknown or unspecified race in the MIMIC-III dataset were included in the others group.

| | **Northwestern Medicine** | **Community Hospitals of YNH** | **MIMIC-III** |
|---|---|---|---|
| **All** | 11,513 | 139 | 146 |
| **Female** | 5,386 (46.7) | 53 (38.1) | 51 (34.3) |
| **Male** | 6,127 (53.3) | 86 (61.9) | 96 (65.7) |
| **Age < 65 years** | 3,727 (32.4) | 29 (20.8) | 38 (25.4) |
| **Age >= 65 years** | 7,786 (67.6) | 110 (79.2) | 109 (74.6) |
| **Race** | | | |
| **White** | 7,460 (64.7) | 108 (77.6) | 104 (71.2) |
| **Black** | 2,466 (21.4) | 14 (10.0) | 15 (10.2) |
| **Hispanic** | 663 (5.7) | 12 (8.6) | 2 (1.3) |
| **Others** | 954 (8.2) | 5 (3.5) | 25 (17.1) * |
| **LVEF < 40%** | 4,678 (40.6) | 70 (50.3) | 74 (50.6) |
| **LVEF >= 40%** | 6,835 (59.4) | 69 (49.7) | 72 (49.4) |

**Table S5. Model Performance Across Quintiles of Note Length.** Data represent model performance metrics in detecting HFrEF across quintiles of word count in discharge summaries from the held-out test set at Yale New Haven Hospital. Abbreviations: AUROC, area under receiver operating characteristic curve; AUPRC, area under precision recall curve; PPV, positive predictive value; NPV, negative predictive value.

| | **AUROC** | **AUPRC** | **Sensitivity** | **Specificity** | **PPV** | **NPV** |
|---|---|---|---|---|---|---|
| **Quintile 1** | 0.96 | 0.94 | 0.90 | 0.92 | 0.94 | 0.88 |
| **Quintile 2** | 0.97 | 0.95 | 0.86 | 0.95 | 0.89 | 0.93 |
| **Quintile 3** | 0.97 | 0.96 | 0.88 | 0.94 | 0.90 | 0.92 |
| **Quintile 4** | 0.98 | 0.99 | 0.93 | 0.92 | 0.90 | 0.94 |
| **Quintile 5** | 0.97 | 0.98 | 0.91 | 0.97 | 0.90 | 0.98 |

**Table S6. Confusion Matrix for Disease Classification.** Table represents confusion matrix of disease classification based on model predictions and ICD-based diagnosis codes on the held-out test set at Yale New Haven Hospital, with echocardiography measurement of left ventricular ejection fraction as the gold standard. Ejection fraction less than 40% was considered as low EF. Abbreviations: ICD, international classification of disease; EF, ejection fraction.

| | | | ICD | |
|---|---|---|---|---|
| | | **Model** | **Normal EF** | **Low EF** |
| **Echo** | **Normal EF** | **Normal EF** | 1658 | 277 |
| | | **Low EF** | 93 | 28 |
| | **Low EF** | **Normal EF** | 153 | 46 |
| | | **Low EF** | 1031 | 690 |

**Table S7. Net Reclassification Improvement of Disease Diagnosis.** Table represents reclassification of HFrEF diagnosis comparing model predictions with chart-documented diagnosis of heart failure phenotype. Abbreviations: NRI, net reclassification improvement; SE, standard error.

| | Moving UP | Moving Down | Total |
|---|---|---|---|
| **Event** | 1031 | 46 | 1920 |
| **Non-event** | 93 | 277 | 2056 |
| **NRI** | 60.2% | | |
| **SE** | 1.9% | | |
| **Z-statistic** | 30.9 | | |

**Figure S1. Model Probabilities of Left Ventricular Systolic Dysfunction and Ejection Fraction on Echocardiography.** Natural logarithmic transformation was applied to model predicted probabilities of HFrEF in the held-out test set. A regression line was fitted through the plot to model the association between log-transformed model probabilities and left ventricular ejection fraction (LVEF = -3.8(Log Probability) + 27.4, p-value < 0.001). Abbreviation: LVEF, left ventricular ejection fraction.

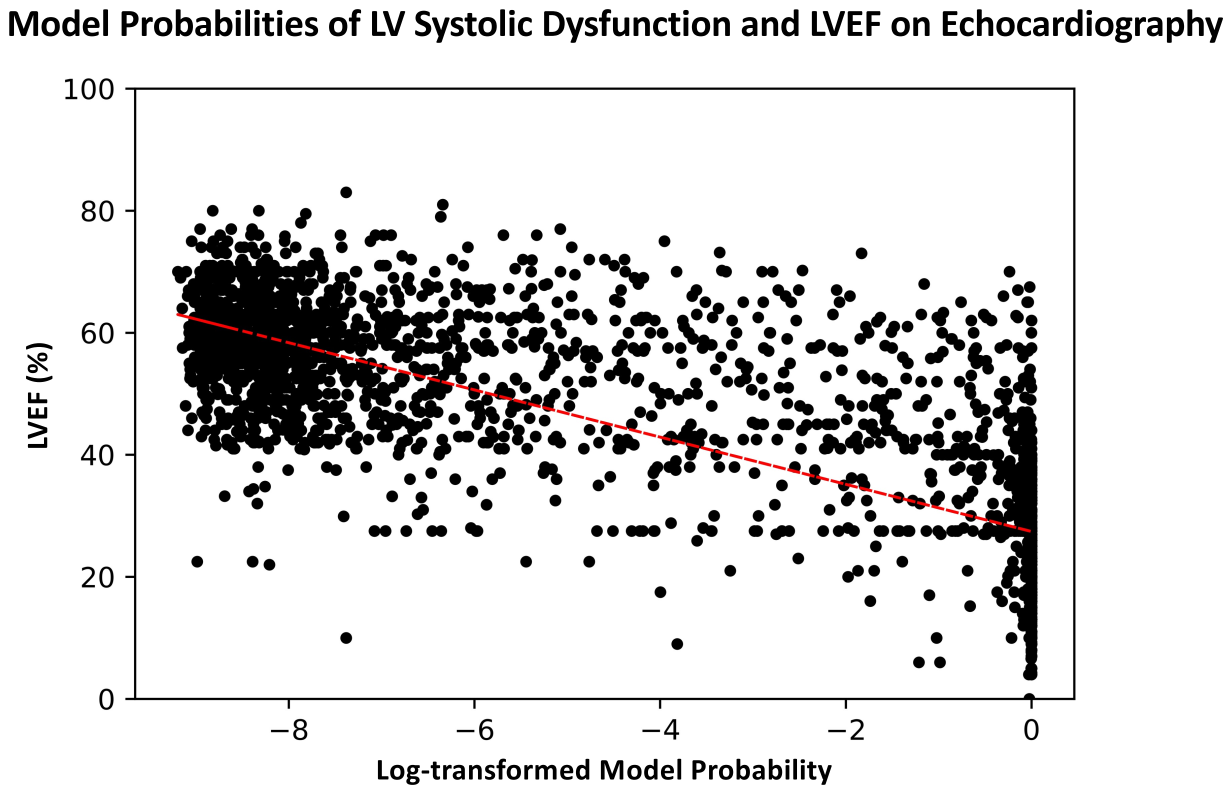

**Figure S2. Model Performance across Notes of Variable Lengths.** The graphs represent receiver operating characteristic (left) and precision-recall (right) curves across quintiles of word count in the discharge summaries from the held-out test set at Yale New Haven Hospital. Abbreviations: ROC, receiver operating characteristic curve; PRC, precision recall curve; AUC, area under curve.

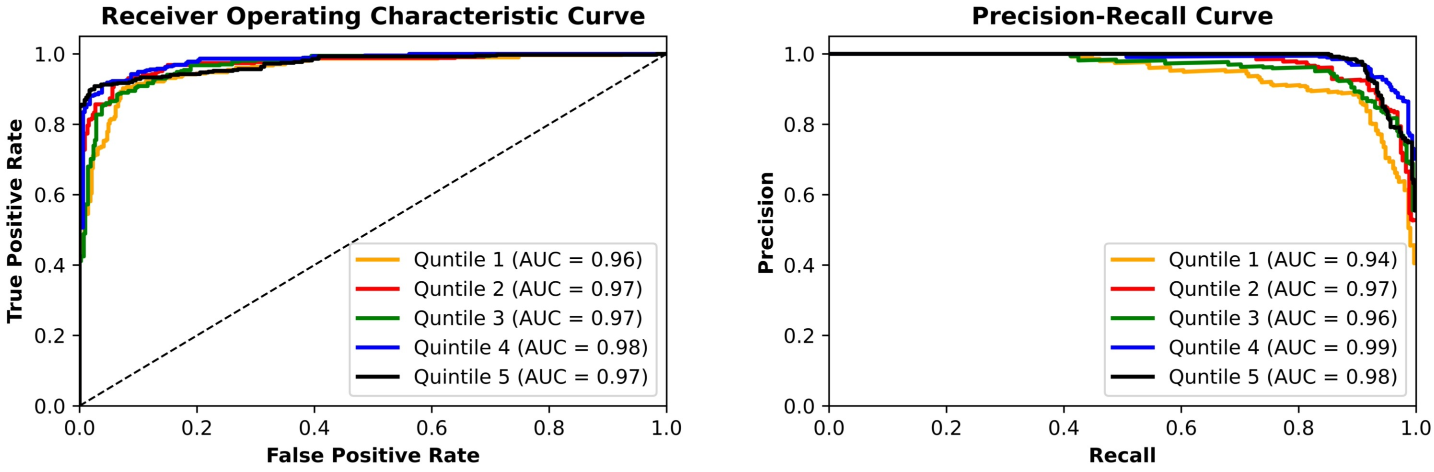

**Figure S3. Model Performance and Ischemic Heart Disease** The graphs represent receiver operating characteristic (top) and precision-recall (bottom) curves among subgroups of individuals with (n=716) and without (n=3,260) ischemic heart disease in the held-out test set at Yale New Haven Hospital. Abbreviations: IHD, ischemic heart disease; ROC, receiver operating characteristic curve; PRC, precision recall curve; AUC, area under curve.

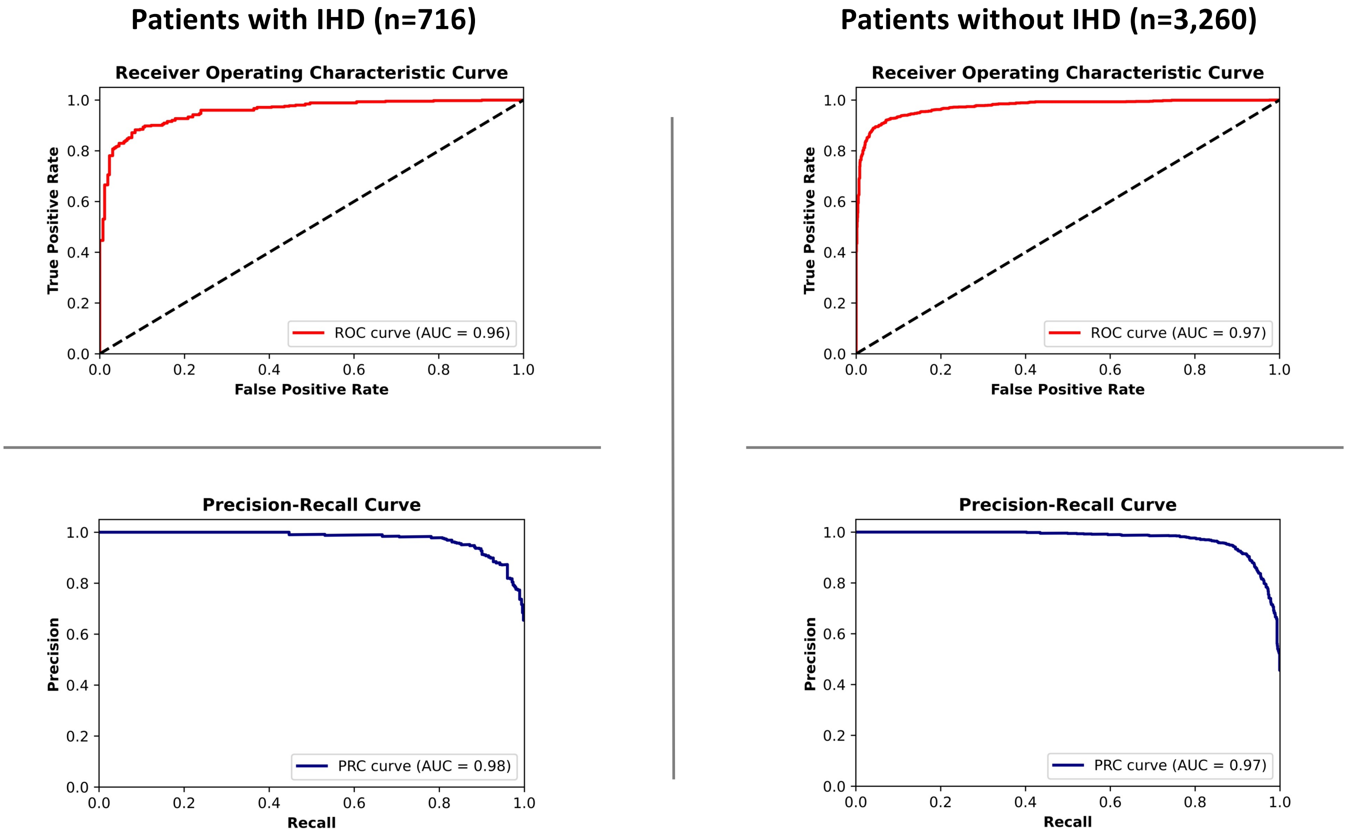

**Figure S4. Model Performance and Atrial Fibrillation/Flutter.** The graphs represent receiver operating characteristic (top) and precision-recall (bottom) curves among subgroups of individuals with (n=1,185) and without (n=2,791) atrial fibrillation and/or atrial flutter in the held-out test set at Yale New Haven Hospital. Abbreviations: Afib, atrial fibrillation; AF, atrial flutter; ROC, receiver operating characteristic curve; PRC, precision recall curve; AUC, area under curve.

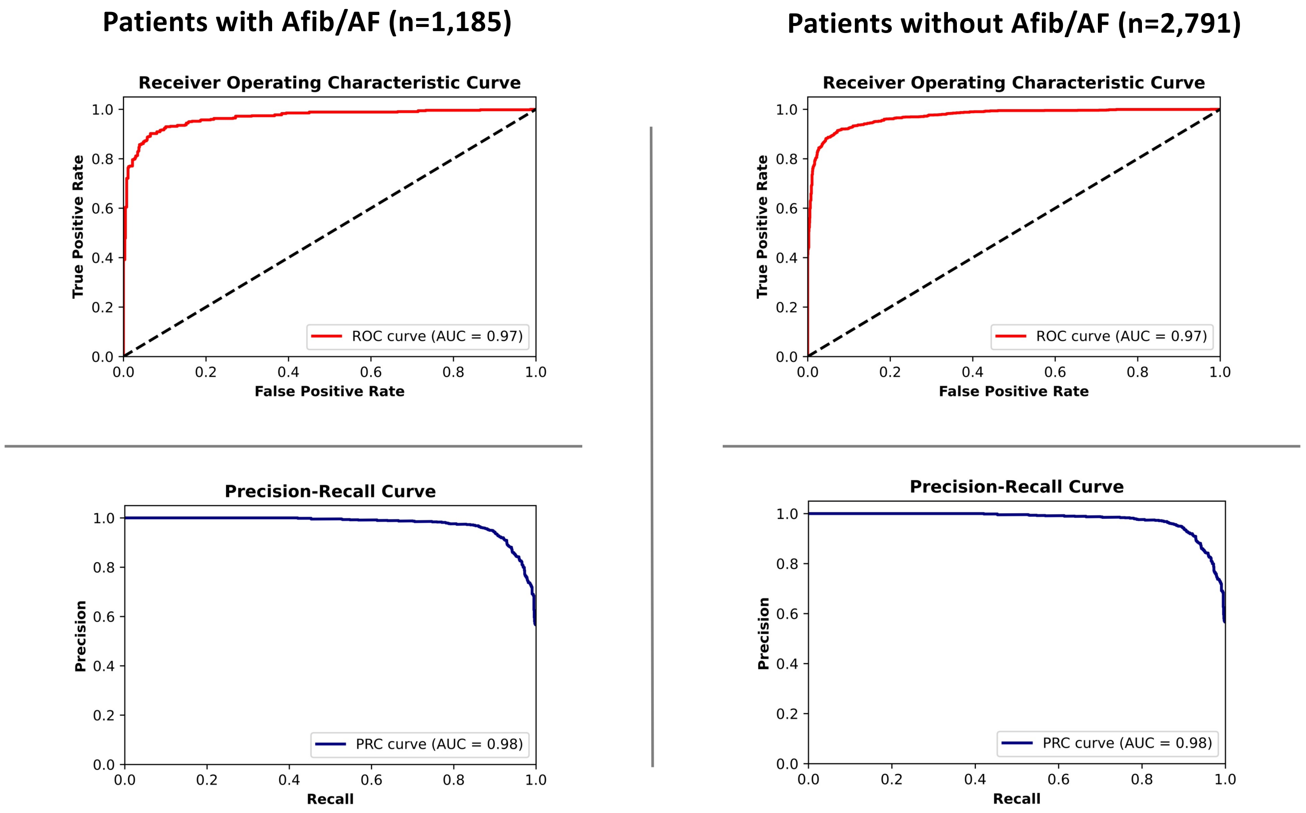

**Figure S5. Model Performance and Right Ventricular Failure.** The graphs represent receiver operating characteristic (top) and precision-recall (bottom) curves among subgroups of individuals with (n=72) and without (n=3,904) right ventricular failure in the held-out test set at Yale New Haven Hospital. Abbreviations: RV, right ventricle; ROC, receiver operating characteristic curve; PRC, precision recall curve; AUC, area under curve.

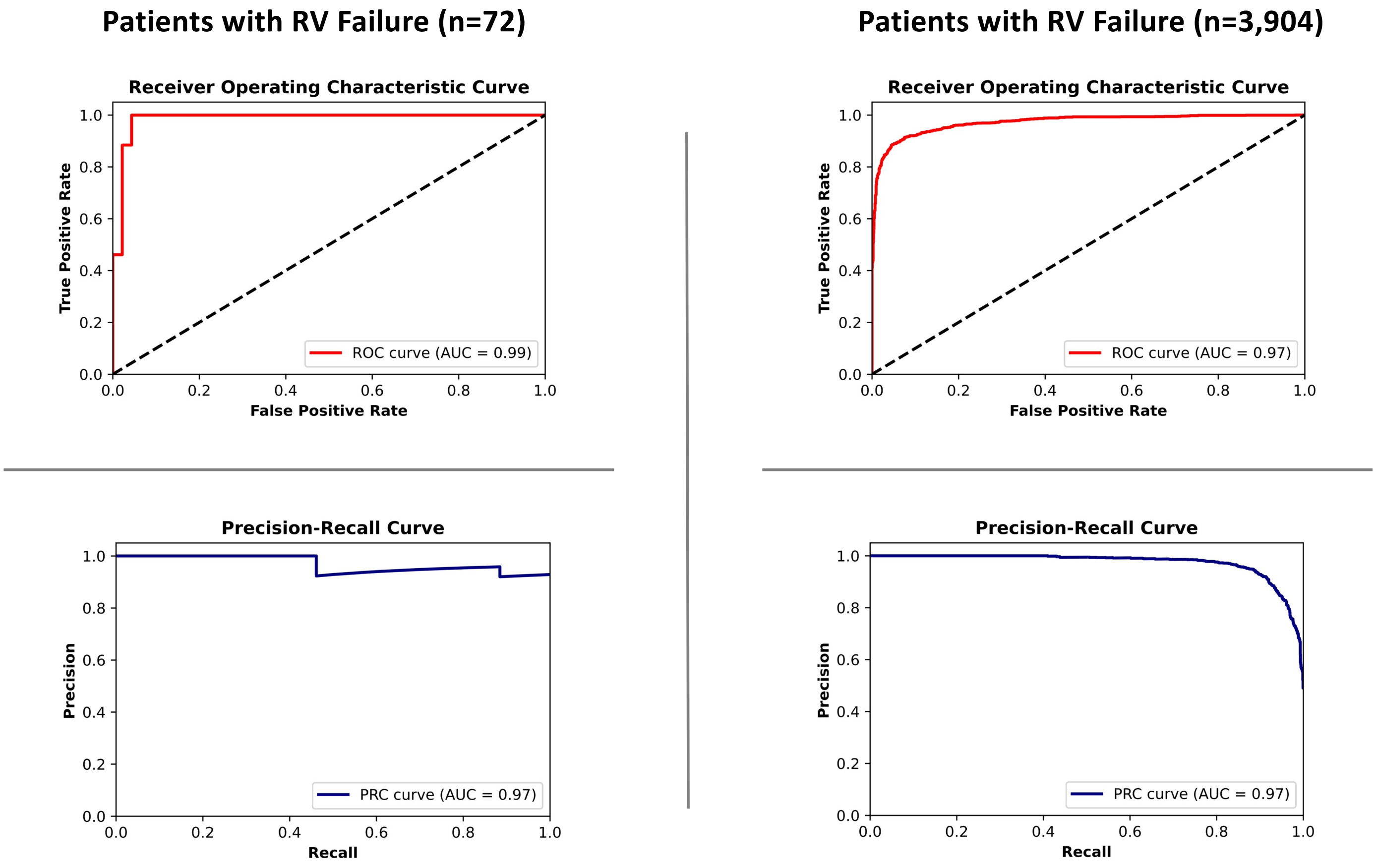
